# Initial Estimates of the Minimal Clinically Important Difference (MCID) for the Neuropathic Pain Symptom Inventory (NPSI): A Systematic Meta-Analysis

**DOI:** 10.1101/2025.01.08.25320237

**Authors:** Alexandra Canori, Rebecca Howard, Jeffrey Bower, David Putrino, Laura Tabacof

## Abstract

**Background:** The Neuropathic Pain Symptom Inventory (NPSI) is a commonly used assessment in neuropathic pain (NP) trials, yet a Minimal Clinically Important Difference (MCID) has not been established. An MCID would enhance the interpretability of NPSI scores, guiding clinicians and researchers in assessing clinically important improvements in NP symptoms. The aim of this study was to calculate an MCID from the available scientific research that used the NPSI.

**Methods:** We conducted a systematic review and meta-analysis of NP trials reporting the NPSI. Four distributional approaches were applied to estimate the MCID: 1) meta-regression on the set of standard deviation (SD) of change scores, 2) meta-regression on the set of baseline SD scores, 3) simple aggregation on the set of SD of change scores, and 4) simple aggregation on the set of baseline SD scores. Only treatment arms within Randomized Controlled Trials (RCTs) were examined for MCID estimation. Control arms were examined separately in a sensitivity analysis using the simple aggregation method for both SD of change and baseline SD sets. Bias for each included study was assessed using the Cochrane tool for quality assessment of randomized controlled trials.

**Results:** 323 trials were examined, 12 were selected for inclusion with a total of 17 treatment arms. The calculated MCID estimates for the NPSI total score (range 1-100) were 6.21 for the SD of change meta-regression and 7.1 for baseline SD meta regression. The MCID values aggregated using simple aggregation methods were 7.95 using pooled SD of change scores, 7.8 using pooled baseline SD. Control arms had a MCID of 8.04 for SD of Change and 8.71 for Baseline SD.

**Conclusion:** This study provides preliminary MCID estimates for the NPSI. Limitations include limited data for NP subtypes, highlighting the need for additional anchor-based and etiology-specific MCID research to refine these estimates. These findings can aid in future NP trial design and the interpretation of results.

## 1 Background

Clinical trials have historically focused on establishing statistical significance as the main measure of clinical success. However, statistical significance does not indicate if the amount of improvement was large enough to be considered clinically meaningful. It is imperative for researchers and clinicians to communicate the clinical importance of their findings to accurately assess its utility to patients. This is especially important when evaluating conditions like neuropathic pain (NP), a multifaceted and chronic disorder that is diverse in presentation and etiology, often causing considerable suffering and distress in patients.

Neuropathic pain is defined by the International Association for the Study of Pain as pain caused by a lesion or disease of the somatosensory nervous system resulting from complex pathophysiological mechanisms involving both peripheral and central nervous system dysfunctions [1]. NP affects roughly 10% of the general population and causes a decrease in quality of life and high healthcare costs, with significant economic burden due to both direct medical expenses and loss of productivity [2,3,4]. NP can either be spontaneous or evoked by stimuli, and presents with negative (e.g. numbness), or positive (e.g. paresthesia) symptoms [5]. To definitively diagnose NP, there must be evidence of damage to the nervous system, through radiological, neurophysiological, or tissue testing, accompanying a neuroanatomically accurate pain pattern. Chronic NP is defined as pain lasting at least 3 months and can be divided into peripheral or central etiologies [6]. Peripheral etiologies include diabetic neuropathy, postherpetic neuralgia, and radiculopathy [7]. Central etiologies include brain injuries, including stroke, and spinal cord injury.

NP treatments span a wide range and include anticonvulsive, antidepressant, or opiate analgesic medications as well as topical medications, surgical procedures, and neuromodulation. Evaluating the effectiveness of a diverse set of new and existing interventions is important to inform clinicians on effective treatment options [8,9,10]. NP can be assessed using standardized questionnaires, clinical examinations, and patient-reported symptoms. Reported symptoms such as burning, tingling, numbness, spontaneous pain, and sensory abnormalities support the diagnosis. Questionnaires can be categorized as screening questionnaires (for identifying patients with NP and distinguishing from non-neuropathic pain) and assessment questionnaires (for measuring symptoms and treatment response) [11].

The Neuropathic Pain Symptom Inventory (NPSI) is an assessment questionnaire that has gained popularity in NP trials due to its capacity to characterize neuropathic symptoms and to monitor treatment effectiveness [12,13]. The survey was developed in 2004 and assesses both the quantitative and qualitative properties of NP as a patient-reported outcome (PRO) [14]. It has been validated in more than 10 different languages for patients with both peripheral and central neuropathic pain [15]. It includes 12 items (10 descriptors of symptoms and 2 items for duration of spontaneous or paroxysmal pain) that categorize pain into 5 subscales of NP: burning pain, evoked pain, pressing pain, paroxysmal pain, and paresthesia/dysesthesia. These subscales can be examined individually, or a total score can be computed that assesses overall NP severity. The NPSI total is calculated by summing the 10 descriptor items for a total score of 0-100, with higher scores indicating greater severity. The NPSI is a relatively new scale that is being used more frequently in trials of NP due to its excellent psychometric properties as well as its ability to assess specific domains of NP with its subscales. PROs like the NPSI align with the FDA’s 2022 guidance on integrating patients’ perspectives and using them as valid scientific evidence for assessing clinically relevant symptoms and improvement resulting from an intervention [16].

The Minimal Clinical Important Difference (MCID) is defined as the smallest difference in the instrument score that patients consider meaningful or beneficial [17]. It is particularly useful when evaluating PROs and maintains a patient-centered approach to evaluating clinical trials, allowing for identification of ‘responders’ to an intervention [18]. The MCID is a value used to assess the clinical relevance of a result that is not captured in the statistical comparison of means [19,15]. While statistical tests assess whether a difference exists, they do not necessarily indicate whether that difference was meaningful to patients. Statistical significance is heavily influenced by sample size, where a larger sample can make even small differences statistically significant [20]. In clinical trials, it is not uncommon to observe statistically significant results that are not of a magnitude that indicates a clinical improvement [21]. An MCID provides a criterion researchers can use to evaluate observed improvements from an intervention and provide a clearer picture of clinical importance.

In the absence of an a-priori MCID value from an external source, the FDA recommends the calculation of the MCID using the anchor method [22,23]. The anchor method is preferred as its estimate of MCID is determined by relating change on the PRO to an established measure of global improvement. Distributionally-based MCIDs are only used in preference to anchor-based when the latter is not available [20,18]. Distributionally-based MCIDs have the disadvantages of being disconnected from any other measure of improvement and they tend to align more with statistical significance, such that a trial with a statistically significant result will often calculate a MCID that is less than their reported change, which does not add much information about clinical importance. This can be remedied somewhat by using MCID estimates that are external to the trial being conducted and are either from earlier pilot trials or from published research. An advantage of distributional methods is that they can often be calculated from reported statistics in published manuscripts, while calculating an MCID based on the anchor method, subject-by-subject data is necessary, and thus, cannot be calculated based on published results data. To estimate an MCID for the NPSI total based on published trials, using a distributional method is necessary.

There are several distributional methods, however most of them involve quantifying the variance of change scores from pre-post intervention. Some of the most common are using the Standard Error of the Measurement (SEM) as the MCID, or by multiplying the Standard Deviation (SD) of change by a value chosen from a range of 0.25 to 0.5. The most common approach is to take ½ of a standard deviation of the change score of a treatment group within a trial [20]. It has been found that ½ a standard deviation corresponds to MCIDs calculated using anchor-based methods within a variety of trials, is highly correlated to using SEM as the MCID, and may represent the limits of human discrimination ability [24]. Smaller fractions, such as ¼ or 1/3 SD, can be used to detect small changes but are vulnerable to measurement error. Thresholds larger than ½ SD produce more conservative MCID estimates, however, since the perceptual threshold corresponds to ½ SD, using these higher thresholds may not accurately measure the *minimum* amount of improvement appropriate for a MCID threshold in most contexts. For this meta-analysis, we have determined that ½ a standard deviation is an appropriate value to use.

As of August 2025, no established MCID exists for the NPSI Total Score. The aim of this meta-analysis was to calculate an MCID from the available scientific research that used the NPSI to measure treatment response. Understanding the amount of change needed in the NSPI score for a clinically important improvement in NP symptoms will help clinicians and scientists to evaluate the effectiveness of previously published interventions and inform future trial design for new NP treatments.

## 2 Methods

### 2.1 Study Design

Clinical trials and randomized controlled trials that evaluated changes in pain using the NPSI in adults over the age of 18 were included. Case reports, abstracts, conference proceedings, reviews, and publications that were unavailable in English were excluded during screening. Studies that used translated versions of the NPSI were considered for inclusion if all questions were a direct translation of the English version, however studies were excluded if questions were not identical across each of the domains. During full-text review, studies were excluded for containing duplicate data, insufficient data to calculate overall NPSI score or change in NPSI score, or missing statistical values. One included study used the validated Thai version of the NPSI, all other included studies used the English version. The exclusion process with these selection criteria is outlined by the PRISMA flowchart in Figure 1. This protocol was developed in accordance with PRISMA guidelines and registered on PROSEPRO (CRD42025649343).

**Figure 1:**
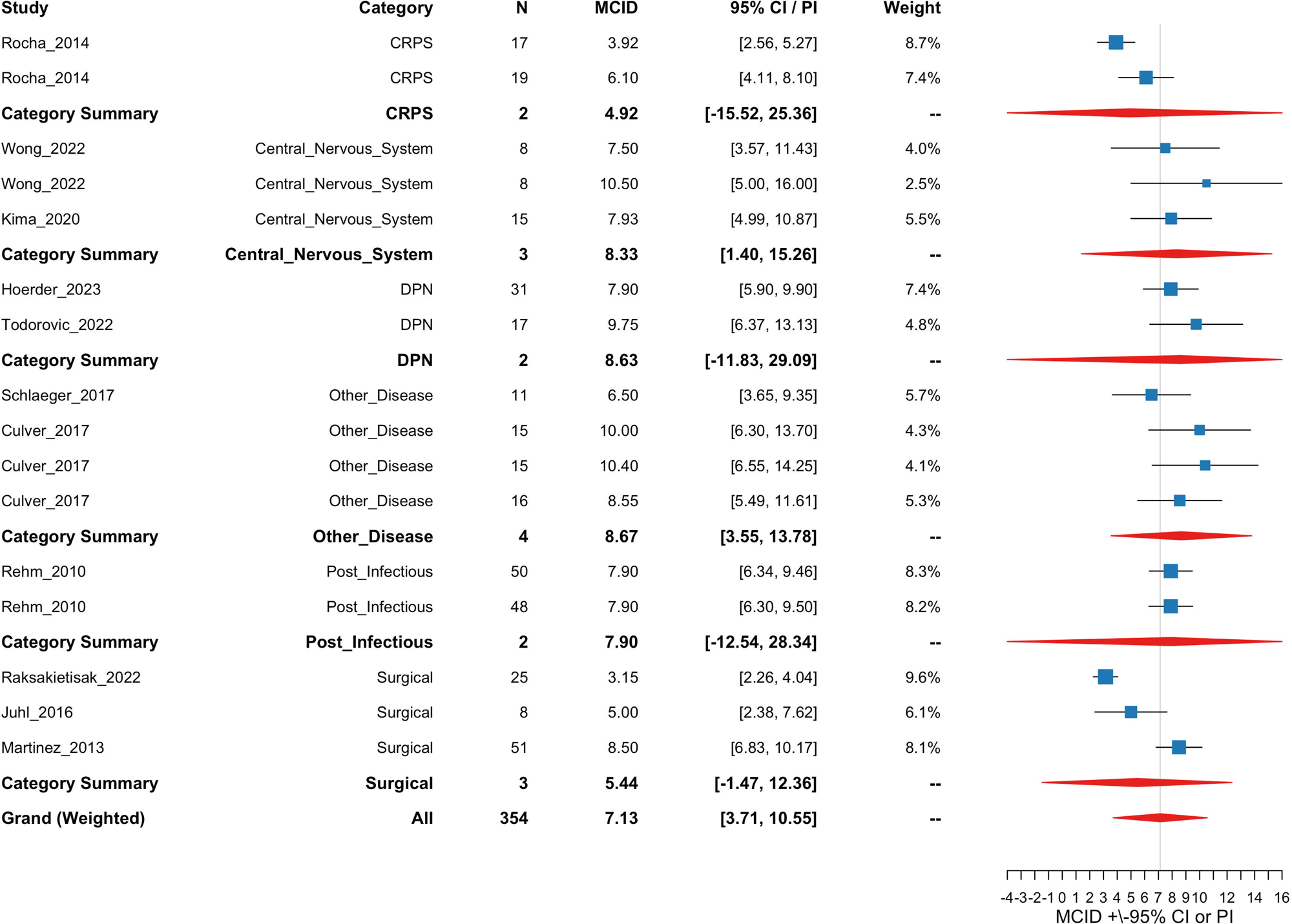
PRISMA Flow Chart

### 2.2 Literature Search

PubMed, Web of Science, and the Cochrane Library databases were systematically searched for all relevant studies. Search dates included studies from 2004, when the NPSI was validated, to February 2025. Unpublished studies were evaluated for inclusion from clinicaltrials.gov. Our search criteria included studies with both significant and non-significant results. A literature search was performed using the following search terms in PudMed, Web of Science, and Cochrane Library, respectively: (“Neuropathic Pain Symptom Inventory”[Title/Abstract] OR “NPSI”[Title/Abstract]) AND (“randomized controlled trial”[Publication Type] OR “clinical trial”[Publication Type]), ((TI=(Neuropathic Pain Symptom Inventory OR NPSI)) AND AB=(Neuropathic Pain Symptom Inventory OR NPSI) AND PY=(2004-2025)), “Neuropathic Pain Symptom Inventory” OR “NPSI” in Title Abstract Keyword - with Publication Year from 2004 to 2025 in Trials. A preliminary screening of titles and abstracts by two independent reviewers was followed by a review of full-text articles, which informed subsequent study selection. Inter-rater agreement was assessed using Cohen’s Kappa, which indicated strong agreement for both title and abstract screening (κ = 0.96) and full-text screening (κ = 0.81). All disagreements were resolved through discussion and consensus was reached. A third reviewer was designated for arbitration purposes, but was not required for this review.

### 2.3 Data Extraction and Quality Assessment

Data extraction was conducted by one reviewer and independently verified by a second reviewer. Two investigators extracted title, author, publication date, study design, pain condition, sample size, mean age, sex, pain intervention, follow-up duration, baseline NPSI score, and follow-up or change in NPSI scores. Data was recorded electronically in a secure, shared spreadsheet. PDF files of all full text studies and available supplementary materials were saved in a shared folder for collective access and reference. Two reviewers independently performed data quality assessment with a third reviewer assigned to resolve disagreements. Methodological quality and confidence of each trial was assessed by reviewing methods of randomization, allocation concealment, blinding, completeness of outcome data, and selective reporting with the Cochrane Risk of Bias (RoB) 2 tool for quality assessment of randomized controlled trials [25]. The GRADE tool was used to assess level of certainty rating.

### 2.4 Study Selection for Analysis

The initial search yielded 341 papers which were assessed for eligibility. A search of trial registries was conducted for unpublished data, however no reports that met data requirements were available. Of the 341 articles, 59 duplicates were excluded, 136 were excluded by title and abstract screening, and 126 were excluded during full-text review (*Figure 1*). The most common reason for exclusion in full-text review was insufficient data to calculate a change in NPSI score. The remaining 20 articles were examined to determine if they reported the required summary statistics or if they were derivable from other reported values. Studies were required to have reported either the SD of change or Baseline SD for the NPSI Total to be included in one of the analyses. In addition, each study was checked to ensure that the NPSI was calculated by convention (sum of 10 items). Within selected trials, only treatment arms were included in the analyses and data from placebo, waitlist, or inactive sham were excluded. After all criteria were assessed, 12 articles were included in the final set for analysis with 17 treatment arms for the analysis (Table 1).

**Table 1:**
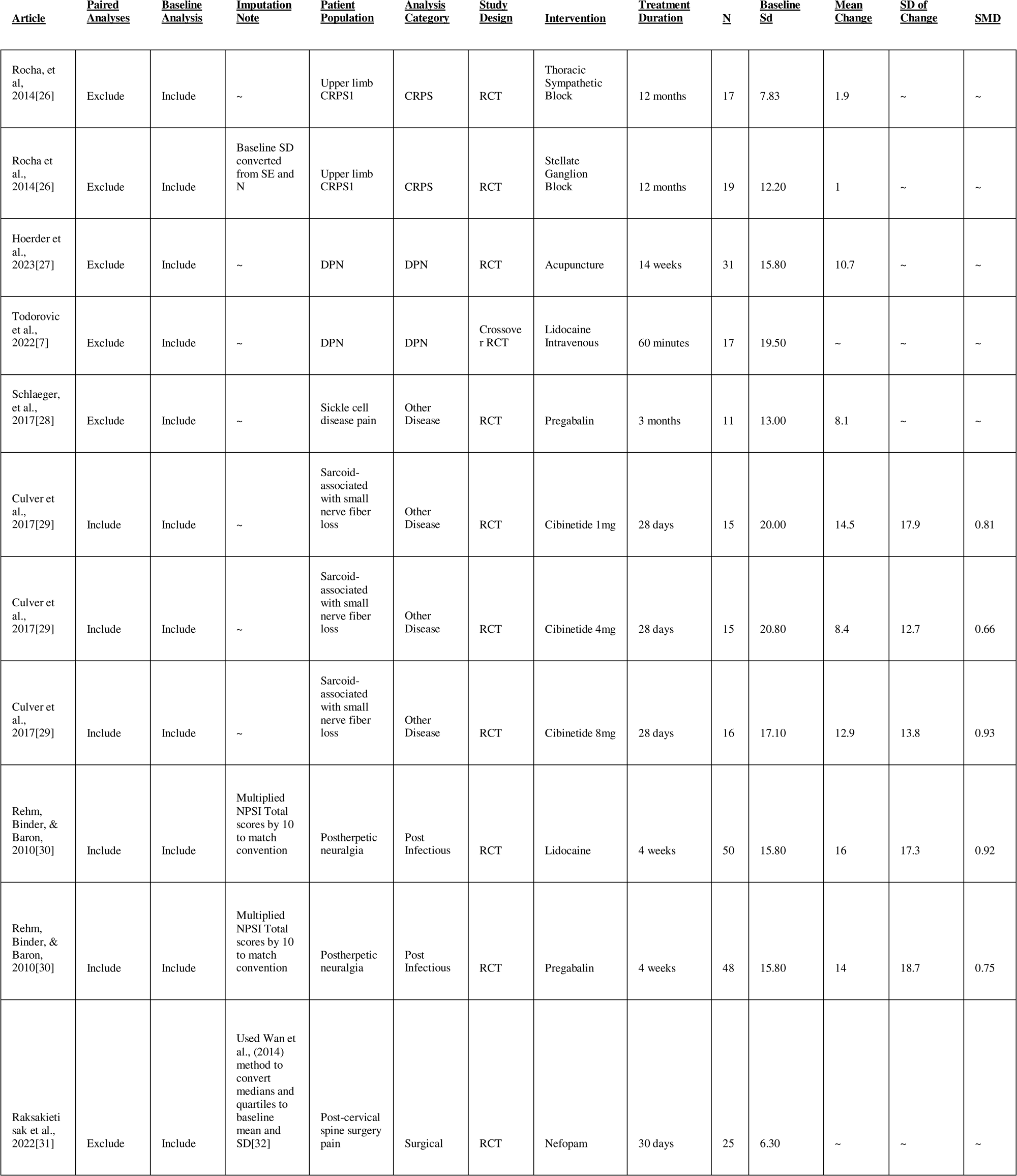
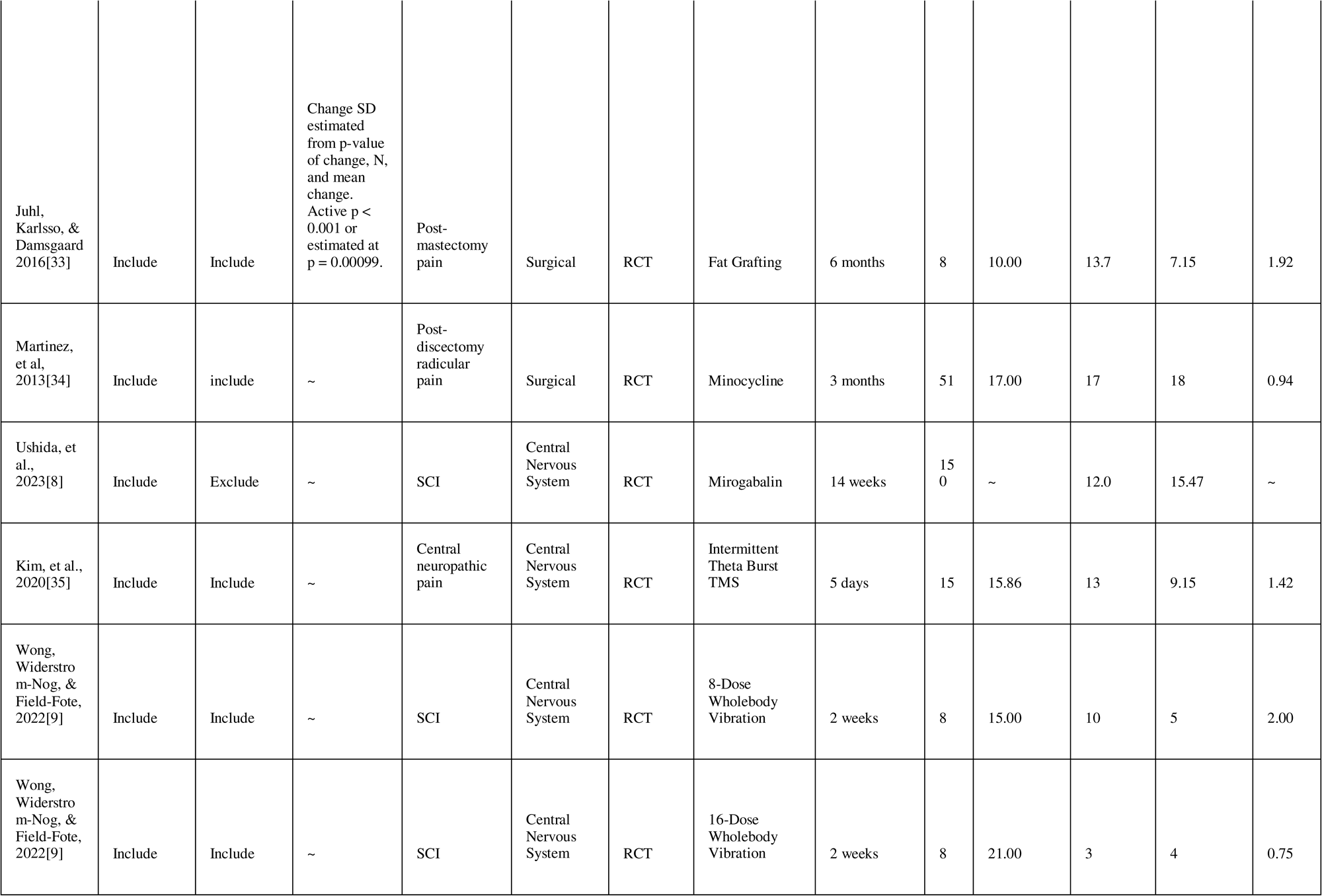
Treatment Arms included in analyses. SMD is the Standardized Mean Difference.

Articles in the final set of studies were categorized by the authors into treatment for 6 etiologies of neuropathic pain to enable sub-set analyses as follows (number of trials in parentheses):

- CRPS: Complex regional pain syndrome type 1 (1)
- DPN: Diabetic peripheral neuropathy (2)
- Other Disease: Sarcoid-associated small nerve fiber loss (1), Sickle Cell Disease pain (1)
- Post Infectious: Postherpetic neuralgia (1)
- Central Nervous System: Brain or spinal cord injury (3)
- Surgical: Post-mastectomy pain (1), Post-cervical spine surgery pain (1), Post-discectomy radicular pain (1)

Table 1 lists the study characteristics for all included treatment arms and reported or derived statistics. The Standardized Mean Difference (Mean-Change/SD-of-Change) was calculated for any trial that that reported Mean-Change and SD-of-Change. Any adjustments necessary to estimate missing values or to correct NPSI scaling are reported in the Imputation Note column in Table 1 and were as follows:

- Rocha et al., 2014 [26]: The baseline SD was calculated using the reported Standard Error of the Mean (SE) and the sample size (N).
- Rehm, Binder, & Baron, 2010[30]: The NPSI total score was multiplied by 10 to bring it from 0-10 to a 0-100 score range.
- Raksakietisak et al., 2022[31]: Reported baseline medians and quartiles were converted into the baseline SD using the method presented in Wan et al., (2014) [32]
- Juhl, Karlsso, & Damsgaard 2016[33]: The active arm reported a p value of < 0.001 for the improvement of the active arm over baseline. We estimated this to be a value p = 0.00099 and used the cumulative inverse distribution function to derive the T value for the test of mean change > 0. From the T value the SE of change could be estimated by dividing the mean change by the T value. Finally, the SD of change was calculated by multiplying the SE of change by the square root of the N.

There was a heterogeneous set of treatments and treatment durations in the sample of selected study arms. The interventions included pharmacological, nerve block, alternative, neuromodulatory, and reconstructive treatments (see Table 1, Intervention column). Treatment durations ranged from 1-hour single treatments to multiple dose regimens of up to 12 months. The selected interventions were all from successful trials, where at least one of the treatment arms was statistically different from control, thus the measures of change and variability reflect those of a successful treatment. The inclusion of diverse interventions and treatments lengths into the analyses limits the ability to produce an aggregated MCID for any single treatment type. The interpretation of the resulting MCID across this sample can be interpreted as the MCID for successful treatments for the NPSI total across a diverse range of NP etiologies. The control arms excluded from the main analysis were not intended as treatments. They were excluded from the primary analyses due the conceptual distinction between treatments intended to produce benefit and those intended as a control. Control arms will be examined in a sensitivity analysis using simple aggregation methods for comparison. Meta-analysis will not be performed on the control arms due to the low number of control arms in this sample. Control arms are presented in Table 2 and follow the same formatting and imputation rules as Table 1

**Table 2:**
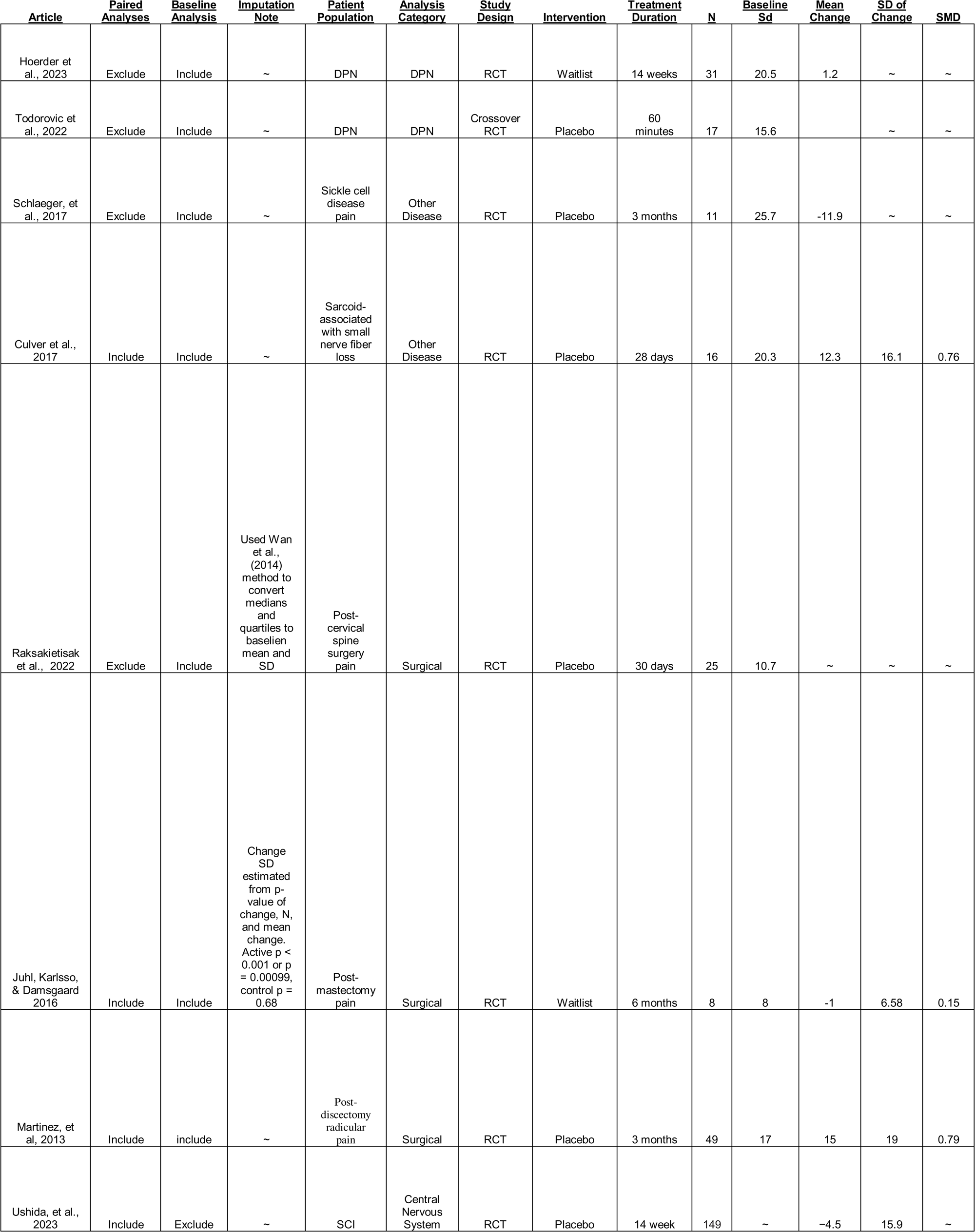
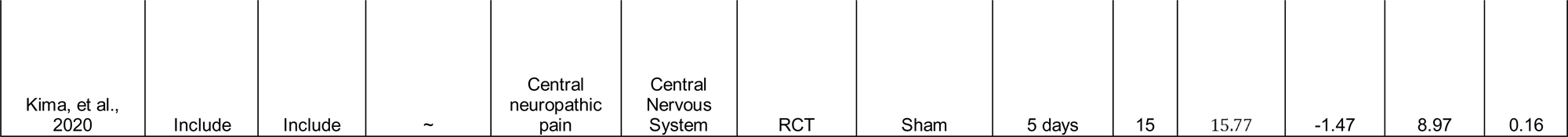
Control Arms included in sensitivity analyses. SMD is the Standardized Mean Difference.

### 2.5 Analyses

Random-effects meta-analysis techniques were adapted to estimate a pooled standard deviation from a set of reported standard deviations, from which the estimates of the effect and confidence intervals which were then divided by 2 to yield the estimated MCID for the NPSI. In a typical meta-analysis, standard deviations from individual studies are used to compute study weights and to estimate heterogeneity. However, when the SD itself is the outcome of interest, it cannot be used to weight its own estimation, and a different variance metric is required.

To estimate the variance across the sample of SDs, we applied the delta method, a well-established statistical technique used to approximate the variance of nonlinear transformations of estimators. Specifically, we used the following approximation (Equation 1):

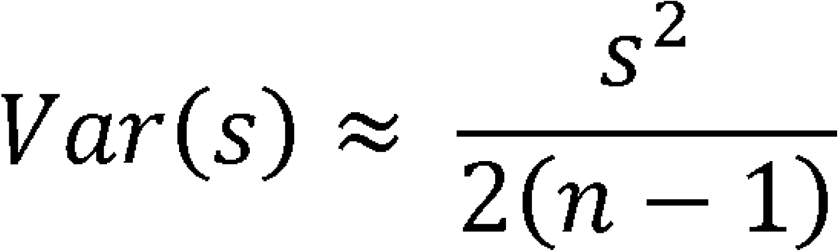

Where s is the standard deviation and n is the number of participants. This formula results from a first-order Taylor series expansion of the square root of the sample variance and assumes the sample SD is approximately normally distributed with moderate skew and kurtosis. The delta method was chosen for its balance of computational simplicity, ease of interpretation, and robust performance.

Alternative approaches were considered, including log-transforming the SD before modeling and estimating the variance of SD based on a chi-squared distribution. However, these approaches were either less interpretable in the context of meta-analytic modeling or required assumptions that were less appropriate given the structure of our data. The delta method was selected as a well justified and empirically supported solution for modeling variance in a meta-analysis of standard deviations. Similar approximations have been used in prior methodological work on estimating sample statistics from summary data, such as in Wan et al. (2014), who used a related formula to estimate standard deviations from medians and interquartile ranges in meta-analyses [32].

A random-effect meta-regression approach was used to model study weights, measures of heterogeneity, and pooled estimates. We fit 2 separate models that used either SD of change or baseline SD score. For the analyses of the first model, we suggested a-priori that the SD of change is the most appropriate statistic to base a MCID upon as it is directly associated with the distribution of change scores resulting from the intervention. However, there is a limited amount of published data that reports the SD of change. Clinical trials almost always report baseline SD. While this source of variance does not capture any aspect of change resulting from the intervention, baseline SD captures variance inherent in the PRO. It has been shown to be highly predictive of the SD of change, and is an appropriate approximation for the calculation of an MCID [20,17,24,36].

In a model selection step, we compared an intercept only model with a model that included etiology category as a moderator (also understood as a categorical variable) by testing the differences in log-likelihood (LL) [ΔLL = 2(LL_reduced_ – LL_full_)] between models using a χ^2^ distribution. In both cases, the model with etiology as a categorical moderator variable explained a significantly greater proportion of the variance (p < 0.05).

Meta-regression was performed in R [37] using “rma.uni” function within the “metafor” package [38]. The function fit the models with a general equation of the form (Equation 2):

Where:

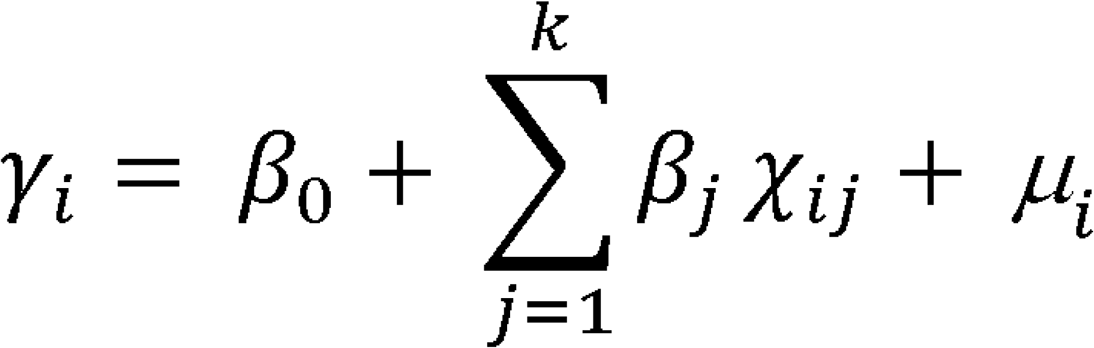

- γ_i_:Summary effect for study arm i. The summary effect will be the Pooled SD in our context.
- β_0_: The model intercept (reference group)
- χ_ij_: Dummy coded moderator etiology categories
- μ_i_: The random effect for each study arm, i. Normally distributed with a SD of Tau ( 2)

The model calculated the weighted SD estimates for each study arm and a pooled SD estimate for each etiology category using a random-effects meta-regression model. The model used REML estimation to fit the data. The grand pooled estimate was computed as the inverse-variance weighted average of the predicted SDs across all study arms. Confidence intervals (CIs) for summary estimates were calculated using the standard error of the weighted mean and the normal approximation. Prediction intervals (PIs) were calculated by incorporating the between-study variance (τ^2^) and the standard error, using a t-distribution based critical value. Model statistics including the estimated between study heterogeneity τ^2^, the proportion of variance due to heterogeneity *I^2^*, and the amount of heterogeneity explained by the moderators *R^2^.* A Q-test for heterogeneity was also performed.

In addition to the meta-regression approach, a simple aggregation method was used for comparison. This method was used by Watt et al. (2021), and first described by Furukawa et al., (2006), to calculate distributionally based MCIDs for the clinical rating scale MMSE and ADAS-Cog [39, 21]. This method calculates a pooled SD across the selected study arms by weighing each arm’s variance by the arm’s sample size (N) (see Equation 3), such that arms with greater numbers of participants are weighted more heavily. The pooled SD formula is typically applied to combine the SD from 2parallel arms in a Randomized Controlled Trial (RTC), however, it can be extended to incorporate any number of treatment arms.

A pooled standard deviation was either calculated across all treatment arms or across a categorical subset using the following formula (Equation 3):

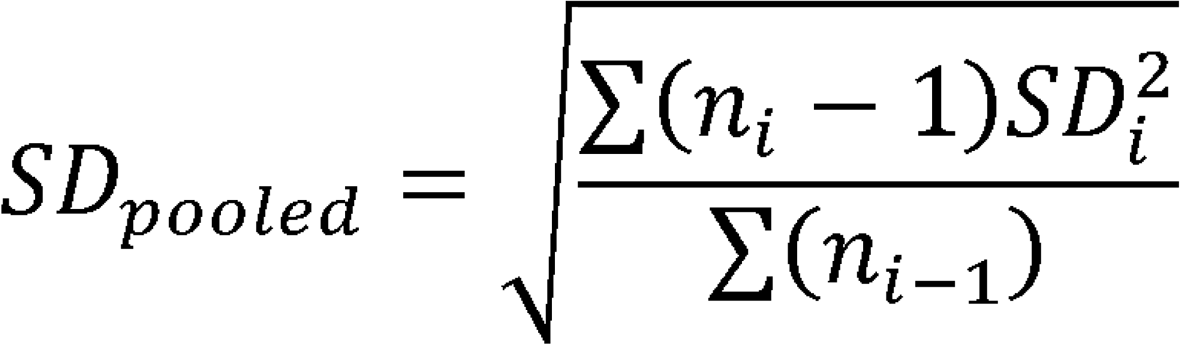

Where *n_i_* is the number of participants and *SD*_i_ is the standard deviation from each treatment arm. Once the SD_pooled_ was calculated, the MCID was calculated as half of the SD_pooled_ Value (SD_pooled_ *0.5). This analysis method was used to calculate MCIDs based on both the paired SD as well as the Baseline SD values.

A sensitivity analysis using equation 3 was performed on the control arms for the paired SD and Baseline SD sets. Control arm data is presented in Table XX below:

Table 2 shows the control arms from the set of studies. Control arms were categorized the same as the treatment arms. Imputation methods are noted on the table in the “Imputation Notes” column and are identical to the methods described for Table 1.

## 3 Results

### 3.1 Quality Assessment

The quality assessment indicates an overall low risk of bias in 75% of the 12 included articles. These findings are highlighted in Figures 2 and 3. In the included studies, level of certainty assessed by GRADE criteria was 67% high, 25% moderate, and 8% low. Figure 2 shows a visual summary of the quality assessment analysis and Figure 3 shows a more detailed quality assessment for each of the 12 analyzed articles.

**Figure 2:**
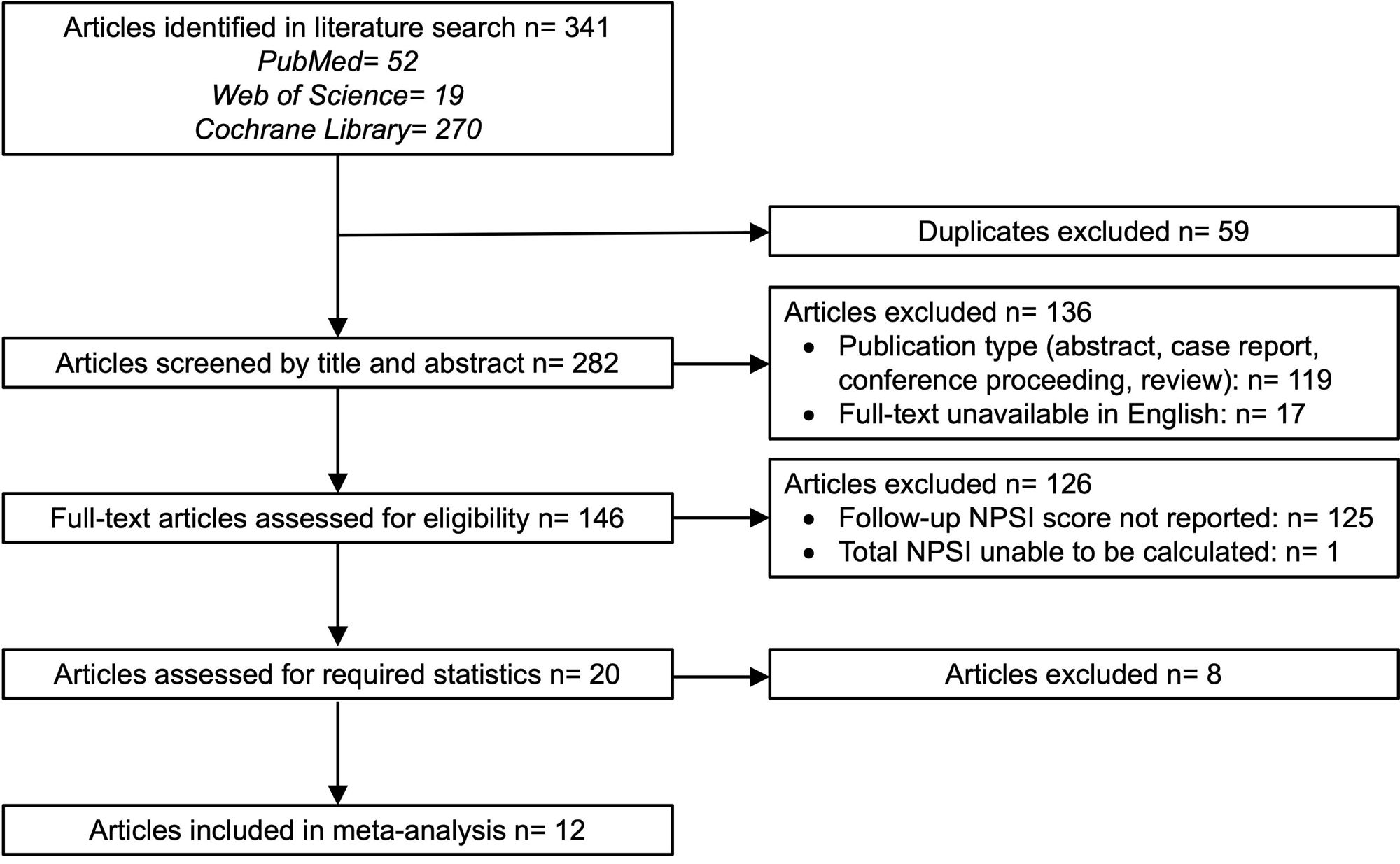
Summary of quality assessment analysis

**Figure 3:**
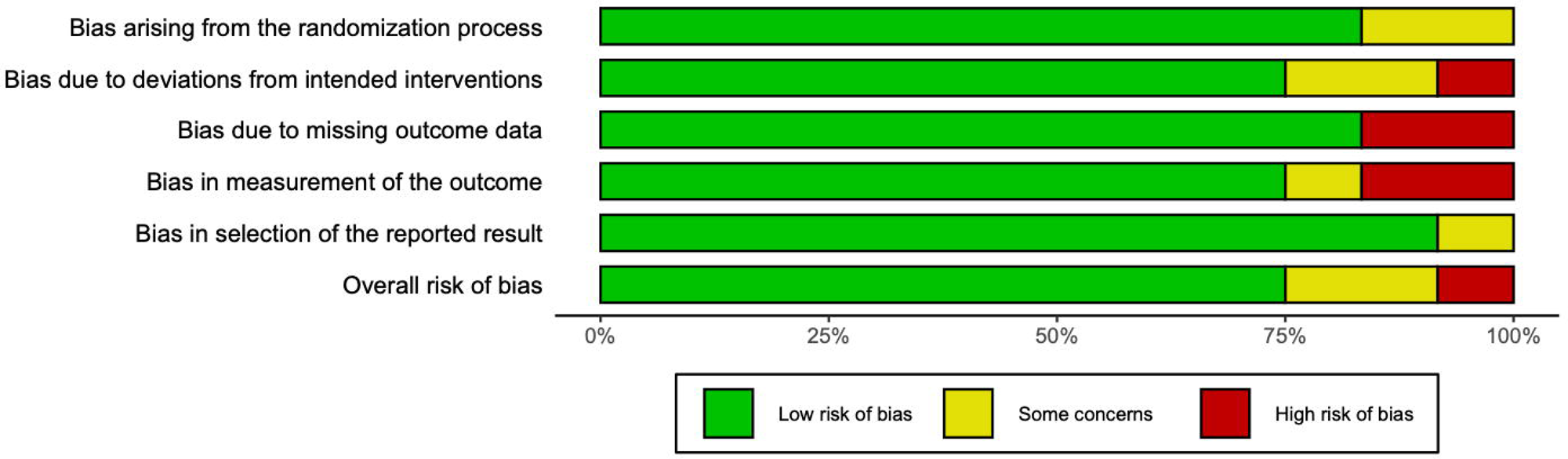
Overview of quality assessment analysis for 12 articles included in this meta-analysis

### 3.2 Meta-Regression – SD of Change

The results for the meta-regression on SD of change indicate the overall pooled estimate of the standard deviation was **12.42**, corresponding to an MCID of **6.21** (MCID = SD / 2). The 95% confidence interval (CI) for the MCID was (6.11, 6.3) with a prediction interval of (1.1, 11.32). There were 11 treatment arms included in this analysis.

Between-study heterogeneity was estimated at τ**² = 21.03**, indicating moderate residual heterogeneity. The proportion of total variability due to heterogeneity was **I² = 89.9%**. The moderator explained **R²= 24.37%** of the between-study variance. The Q-test for heterogeneity was significant (Q = 101.2, *p*< 0.001), indicating that there was significant unexplained heterogeneity between study arms.

Figure 4 shows a forest plot of the meta-regression model with study arm estimates, etiology category estimates, and the grand estimate of the MCID. Category and grand summary estimates are represented by diamonds in which the width is the 95% Prediction Interval (PI) Prediction intervals were computed by combining the standard error of the pooled estimate with the estimated between-study variance (τ²), using a critical value from the *t*-distribution to account for sampling uncertainty and heterogeneity across studies. PIs were clipped at the bounds of the X axis if they exceeded a reasonable range for overall plot visibility.

**Figure 4:**
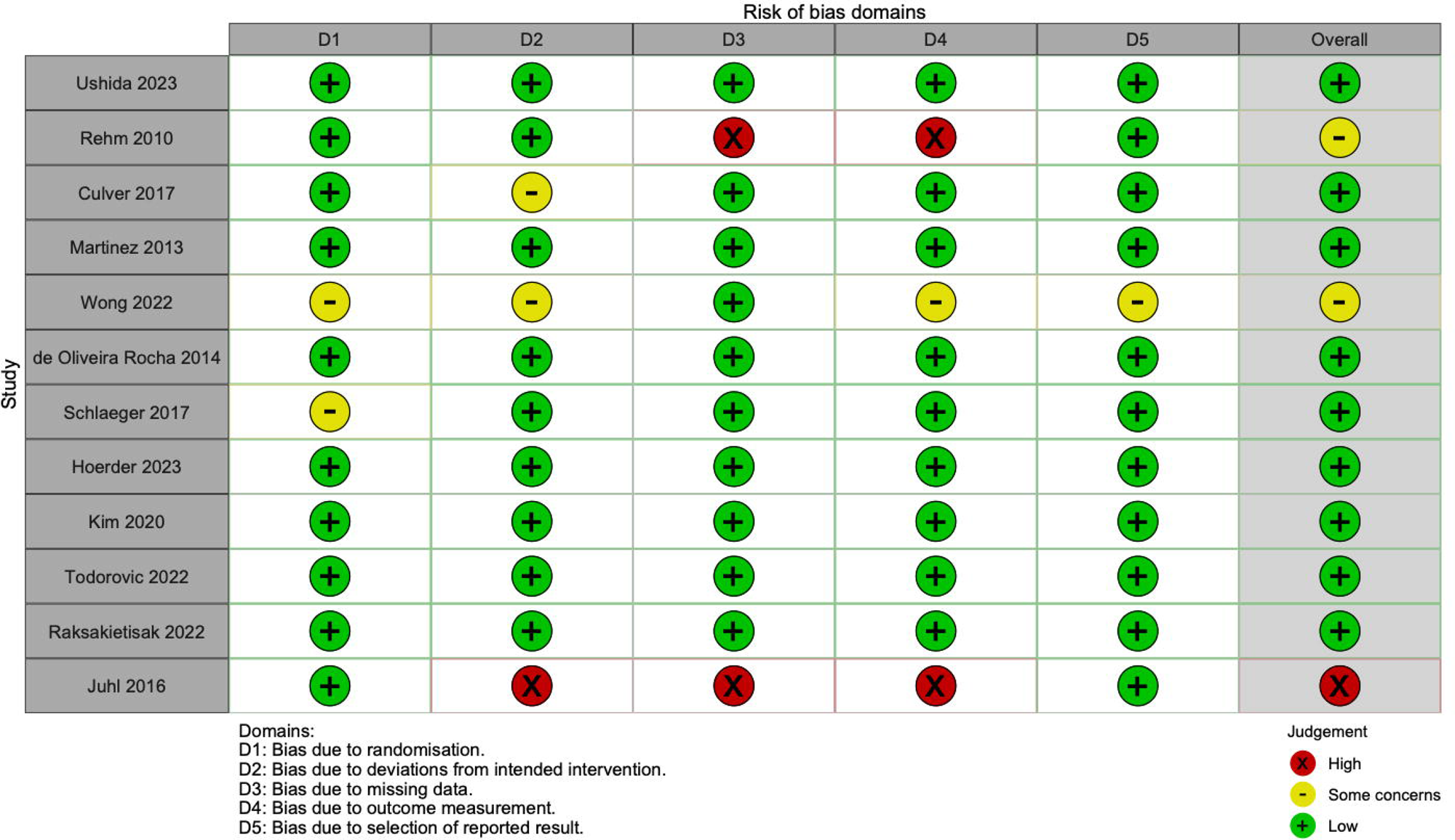
Forest plot of study, etiology category, and grand estimates of MCID for the SD of Change Meta-regression analysis. The X axis is the estimated MCID values. “For 95%CI / PI” column, and for the error bars within the forest plot, CIs are presented for study arms and PIs are presented for category summaries and the grand estimate. Squares are a visual representation of study weights. Diamonds represent PI bounds and are cut off at the limits of the X-Axis for visibility.

Etiology estimates with 95% CIs and 95% PIs are presented in Table 3.

**Table 3:**
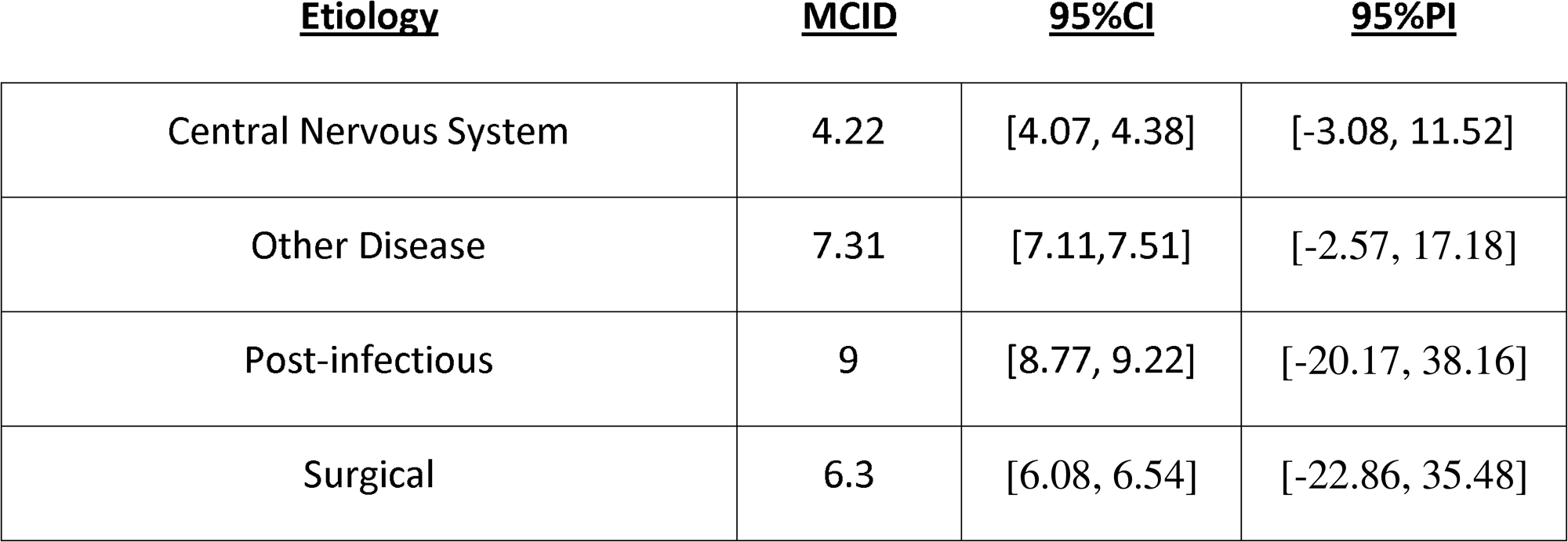
Etiology specific estimates with 95% Cis and 95% PIs for the meta-regression using SD-of-change scores.

Overall, this model had a moderately good fit as indicated by **R² = 24.37%**. However, the was a moderate to large amount of residual heterogeneity, as well as a large proportion of the variance explained by heterogeneity. Estimates of category and grand estimates did have precise CIs, indicating a precise estimation within model, however with very wide PIs. This was most likely due to the small number of study arm in each etiology category.

### 3.3 Meta-Regression – Baseline SD

The results for the meta-regression on Baseline SD indicate the overall pooled estimate of the standard deviation was **14.3**, corresponding to an MCID of **7.1** (MCID = SD / 2). The 95% confidence interval (CI) for the MCID was (7.0, 7.2) with a prediction interval of (3.7, 10.5). There were 16 treatments arms in this analysis.

Between-study heterogeneity was estimated at τ**² = 10.3**, indicating low residual heterogeneity. The proportion of total variability due to heterogeneity was **I² = 66.98%**. The moderator explained **R² = 29.79%** of the between-study variance. The Q-test for heterogeneity was significant (Q = 39.3, df = 10, *p* <0.001), suggesting that there was variability across study arms.

Figure 5 shows a forest plot of the Meta-regression model.

**Figure 5:**
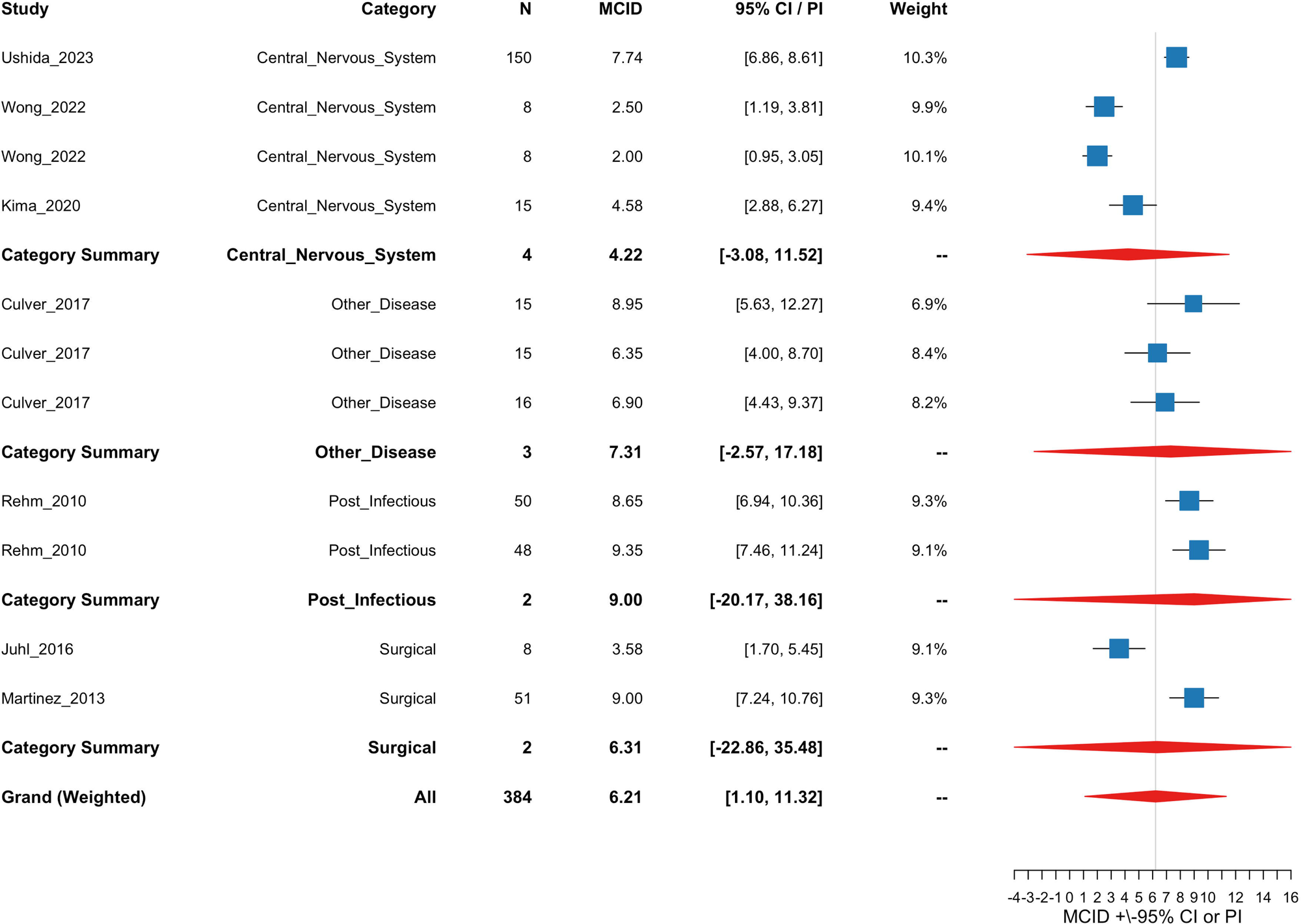
Forest plot of study, etiology category, and grand estimates of MCID for the baseline SD meta-regression analysis. The X axis is the estimated MCID values. “For 95%CI / PI” column, and for the error bars within the forest plot, CIs are presented for study arms and PIs are presented for category summaries and the grand estimate. Squares are a visual representation of study weights. Diamonds represent PI bounds and are cut off at the limits of the X-Axis for visibility.

Etiology estimates with 95%CIs and 95%PIs are shown in Table 4

**Table 4:**
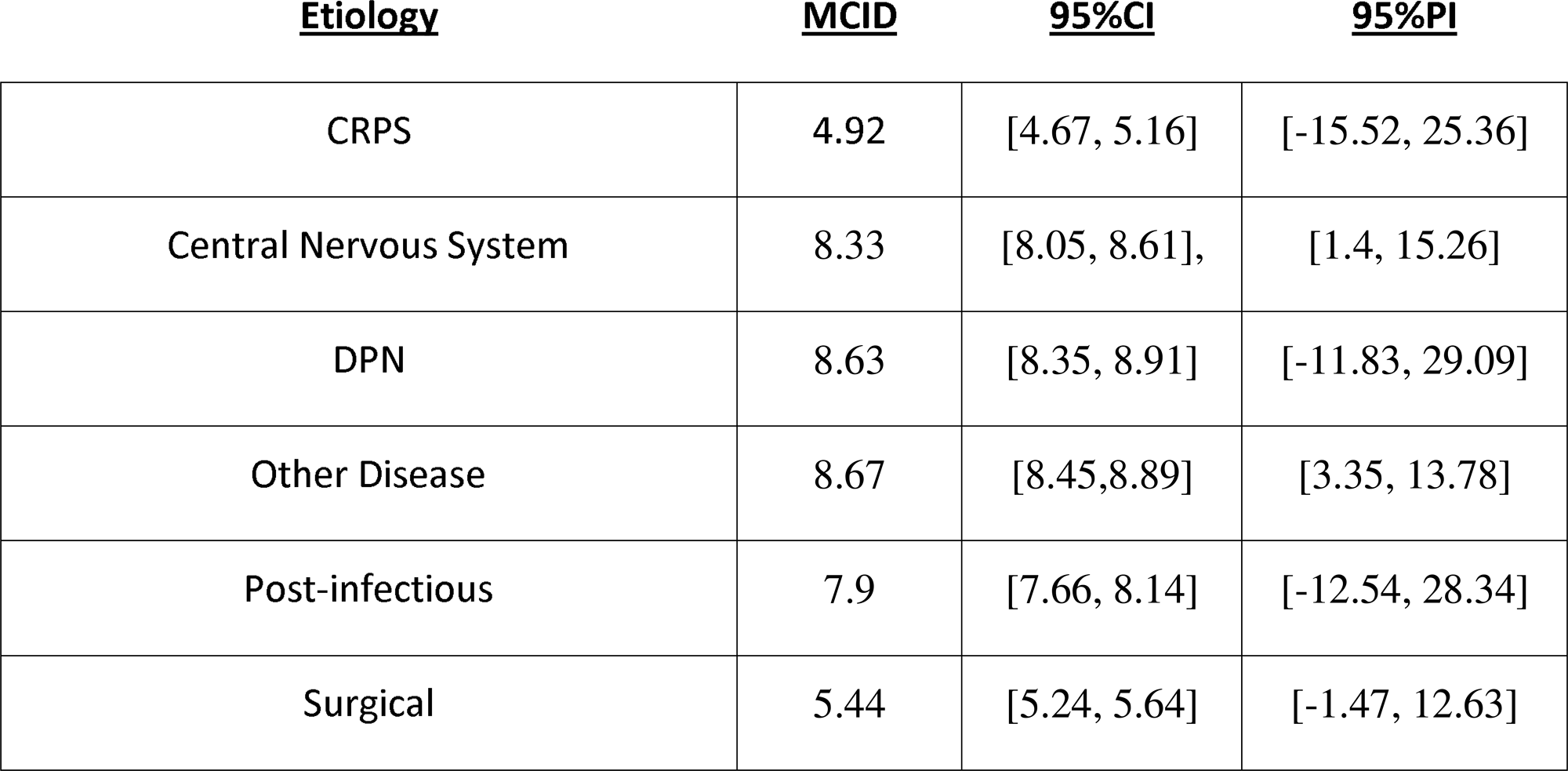
Etiology specific estimates with 95% Cis and 95% PIs for the meta-regression using baseline SD scores.

Overall, this model had a good fit as indicated by **R² = 29.79%** as well as low residual heterogeneity. This model showed that 67% of total variance was due to heterogeneity which is statistically significant as indicated by the Q-test. Differences across categories are the likely source of this heterogeneity. The Baseline SD model has an overall better model fit compared the SD of change meta-regression. Estimates of category and grand estimates have CIs indicating a precise estimation within model. PIs are generally very wide, except within the etiology categories of Central Nervous System and Other Disease as well as for the grand estimate which provided tighter prediction intervals compared to the other estimates.

### 3.4 Pooled SD Method – SD of Change

The results for the SD of Change analyses are presented in Table 5. The results show that the aggregated MCID across all arms that reported SD of change is 7.95 points on the NPSI total. The range of MCIDs reported within each category were 7.24 to 9. There were 11 treatment arms in this analysis.

**Table 5:**
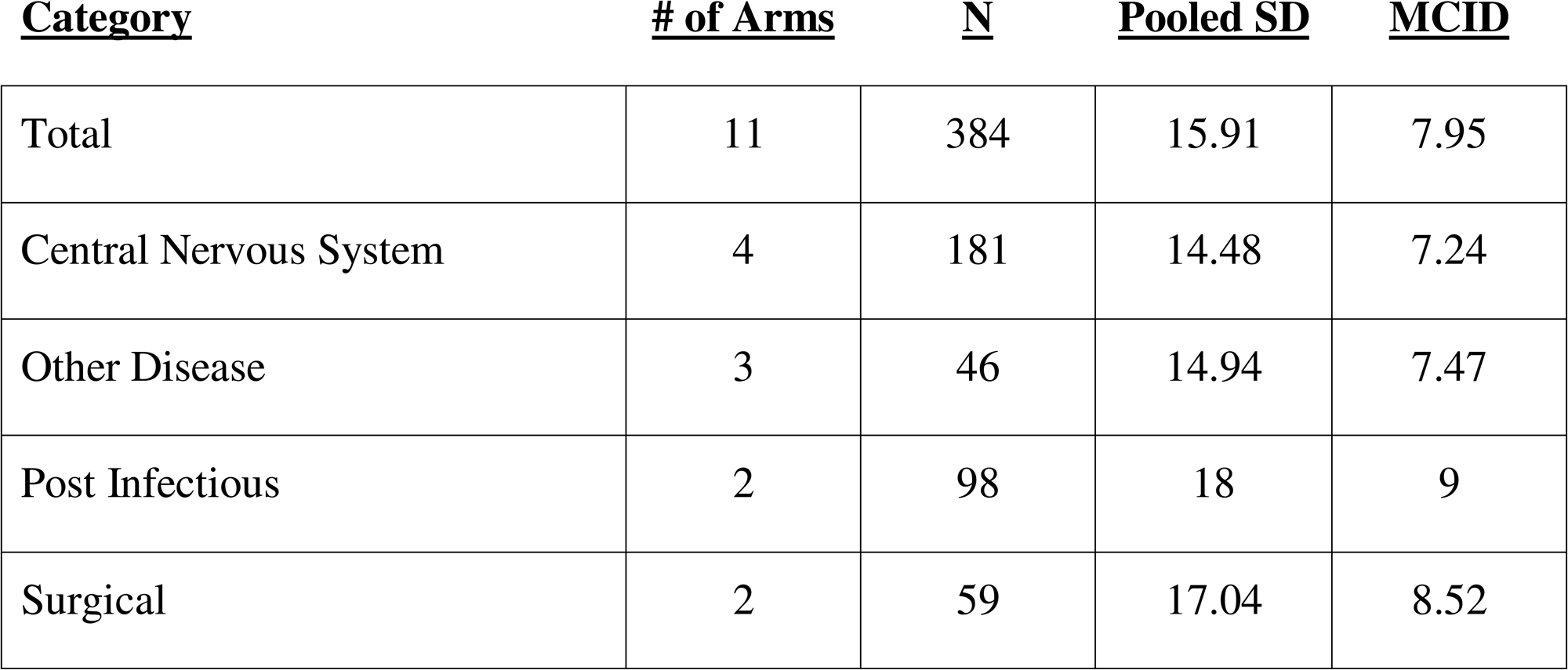
Results of the Pooled-SD-Method – SD-of-Change.

The results for the sensitivity SD of Change analyses on Control arms are presented in Table 6. The results show that the aggregated MCID across the control arms that reported SD of change is 8.04 points on the NPSI total. The range of MCIDs reported within each category were 7.71 to 8.95. There were 5 control arms in this analysis.

**Table 6:**
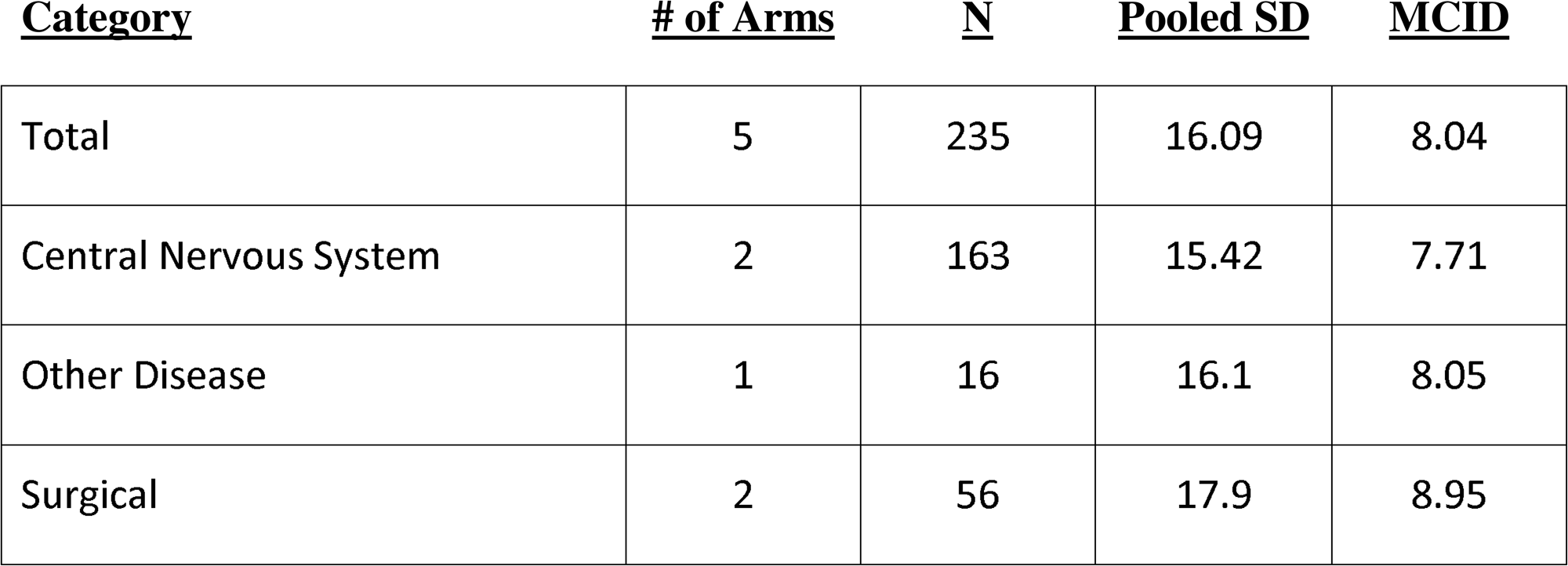
Results of the Pooled-SD-Method for Control Arms – SD-of-Change.

### 3.5 Pooled SD Method – Baseline SD

The results for the Baseline SD analyses are presented in Table 7. The results show that the aggregated MCID across all arms that reported baseline SD is 7.84 points on the NPSI total. The range of MCIDs reported within each category were 5.19 to 9.14. There were 16 arms in this analysis. There were 8 control arms in this analysis.

**Table 7:**
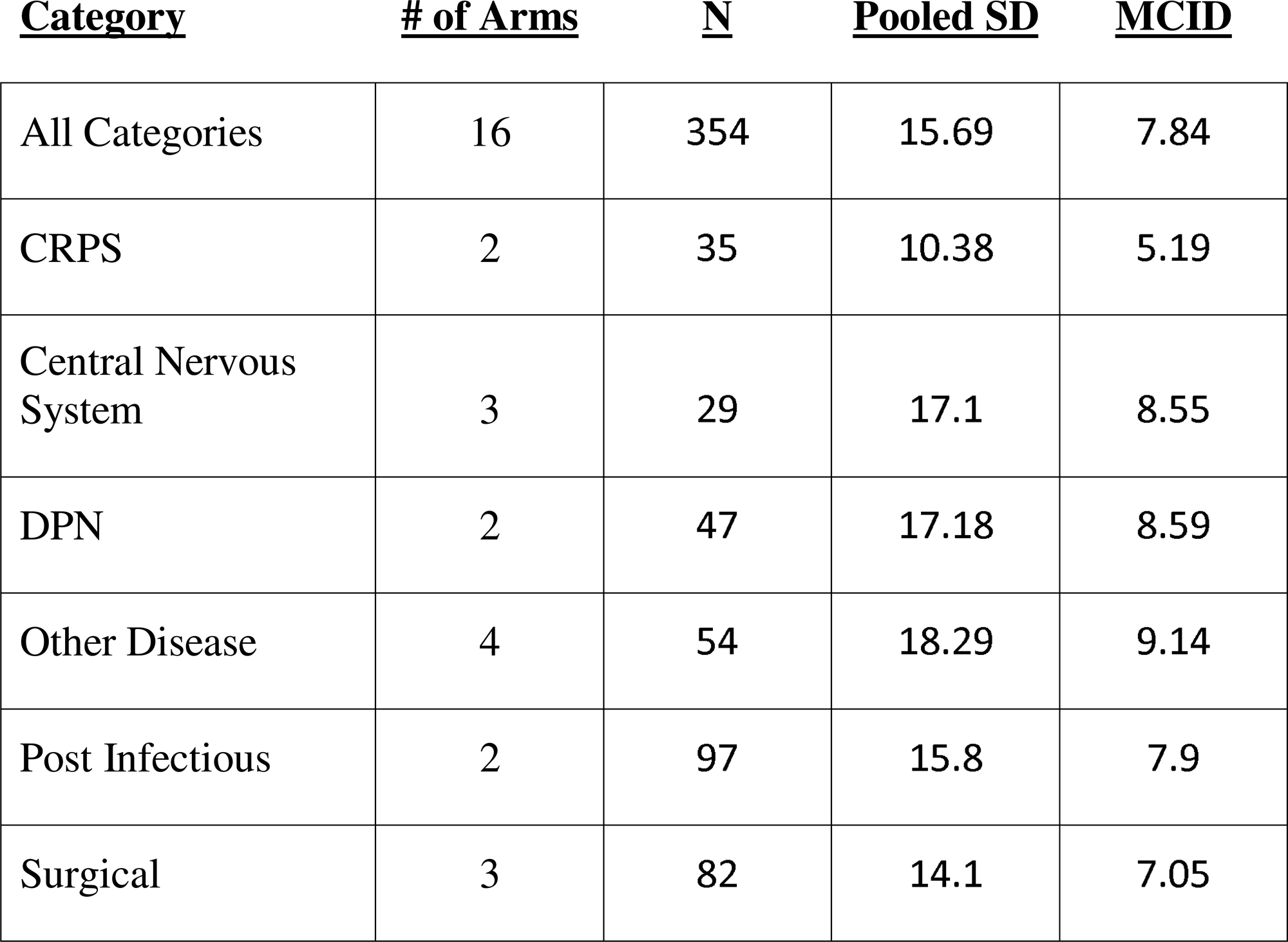
Results of the Pooled-SD-Method – Baseline-SD.

The results for the sensitivity Baseline SD analyses on the Control arms are presented in Table 8. The results show that the aggregated MCID across all arms that reported baseline SD is 8.71 points on the NPSI total. The range of MCIDs reported within each category were 7.35 to 9.37.

**Table 8:**
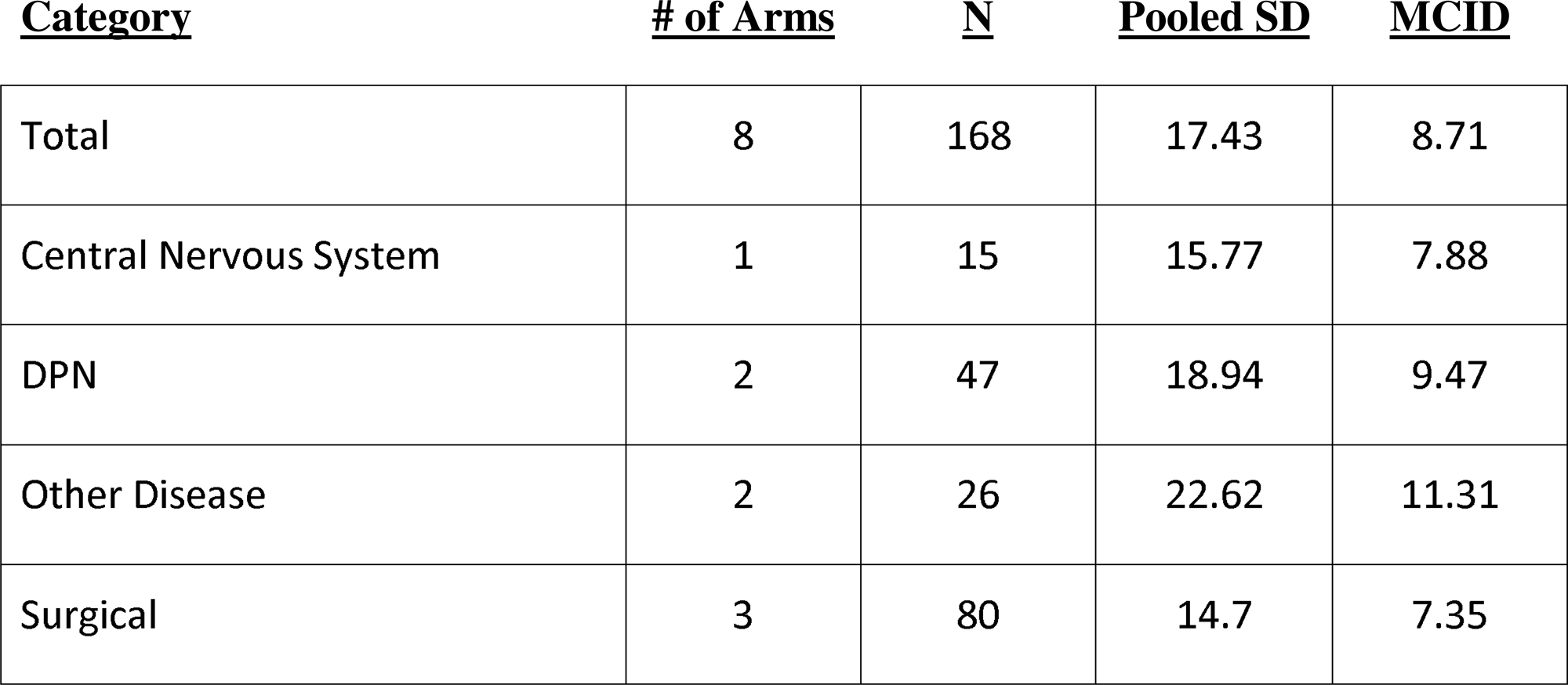
Results of the Pooled-SD-Method for Control Arms – Baseline-SD.

## 4 Discussion

This meta-analysis has calculated two weighed estimates of the MCID for the NPSI total score across a range of NP treatments and etiologies. Two additional estimates were produced using simple aggregation methods. MCID values are critical in clinical research as they provide a threshold to interpret whether observed improvements in symptom scales, such as the NPSI, reflect clinically meaningful changes for patients. These values aid clinicians and researchers in evaluating the practical impact of interventions and designing future trials.

The two meta-regression analyses produced similar estimates of the MCID. The estimates were 6.21 for the SD of change meta-regression and 7.1 the baseline SD meta-regression. Using simple aggregation methods, the MCID estimates were 7.95 for SD of change and 7.84 for Baseline SD. The difference between the largest and smallest MCID is 1.74 points, suggesting that these estimates were highly similar across methods and therefore these results are likely approaching the true value.

Control arms were examined as a sensitivity analysis for both the SD of Change and Baseline SD sets using the simple aggregation method. These arms were excluded from the full analysis on the rationale that they were not intended as real treatments and are conceptually distinct in this regard.However, the control arms were assessed using the NPSI and thus will have similar distributional properties to the active arms. The results of this sensitivity analysis produced an MCID estimate of 8.04 for the SD of change set and an estimate of 8.71from the Baseline SD set. These MCID estimates from the control arms were marginally higher but notable similar to those produced using the active arms. This implies that the variance of the NPSI may be consistent when measuring active or inactive arms and is somewhat independent of the observed treatments effect. Future analyses using distributional methods to estimate MCIDs may consider including the control arms to their analyses as if they are not concerned about the conceptual distinction between active and control. The meta-regression and simple aggregation method for SD of change are based on change scores and their variance, are directly related to the actual measured improvement resulting from an intervention. In contrast, methods based on the baseline SD are disconnected from any measured improvements in a trial, but capture the variance inherent in the NPSI. Based on this reasoning, methods relying on baseline SD are inherently more limited for producing an accurate MCID. However, applying the baseline SD method enabled the inclusion of more data and improved the robustness of MCID estimates across categories. In the case of the meta-regression on Baseline SDs, the fit is clearly improved and produces more accurate estimates of MCID within the model. This was likely due to the inclusion of more trials into the baseline SD compared the SD of change SD sets. In both meta-regressions, CIs were narrow indicating good within model estimates. However, with the exception of Central Nervous System and Other Disease etiology categories, PIs were extremely wide, indicating the prediction accuracy of the model is imprecise.

Examination of the sub-analyses of etiologies produces a different pattern of MCIDs depending on the method used. Table 9 summarizes the MCIDs found across the three methods and shows the average MCID by etiology.

**Table 9:** Aggregated and Mean MCID values across the 4 methods examined.

| <u>Etiology</u> | <u>Meta-</u><br><u>regression</u><br><u>Change SD</u> | <u>Meta-</u><br><u>regression</u><br><u>Baseline SD</u> | <u>Simple</u><br><u>Change SD</u> | <u>Simple</u><br><u>Baseline SD</u> | <u>Mean</u><br><u>MCID</u> |
| --- | --- | --- | --- | --- | --- |
| Central Nervous<br>System | 4.22 | 8.33 | 7.24 | 8.55 | 7.09 |
| Other disease | 7.31 | 8.67 | 7.47 | 9.14 | 8.15 |
| Post-Infectious | 9 | 7.9 | 9 | 7.9 | 8.45 |
| Surgical | 6.3 | 5.44 | 8.52 | 7.05 | 6.83 |
| CRPS | ~ | 4.92 | ~ | 5.19 | 5.06 |
| DPN | ~ | 8.63 | ~ | 8.59 | 8.61 |

Of the four methods, the results in Table 9 show that the Baseline-SD meta-regression and simple aggregation method produce a similar pattern of MCID estimates. The meta-regression and simple aggregation method for SD of change scores differed noticeably from each other in the Central Nervous System and Surgical etiology categories. These were lower overall in the meta-regression, most likely due to the weights in the meta-regression. MCID estimates were more dissimilar between the 2 SD of change methods and the 2 baseline SD methods. This is most likely explained by these analyses being based on different underlying sets of SDs.

The Baseline-SD meta-regression analysis produced the most accurate estimates, and the etiology MCID were of similar values to the other methods. This similarity supports the assumption that baseline SD can be used in place of change-score SD for the calculation of MCID. For this analysis, the baseline SD method was included to increase the number of study arms included in the analysis, as baseline SD is reported more often in Randomized Controlled Trials (RCTs)s than within arm SD of change. CRPS and DPN did not have any published clinical trial that reported a change SD, and thus the baseline method was needed to estimate MCID for these etiologies. Table 9 computes the mean MCID across etiologies. Calculating the mean MCID per etiology produces an aggregated MCID that is likely closer to the true MCID.

The nature of pain may contribute to the variation in MCID between etiologies. Neuropathic pain is multifaceted, with a vast range of mechanisms and factors that contribute to patients’ symptoms, therefore an improvement in certain pain characteristics may affect quality of life more than others. Changes in perceived intensity could affect the range, and thus the variance, of reported total pain scores in PROs which in turn would modulate an MCID based on distributional methods. For example, paroxysmal pain that feels like stabbing or electric shocks may be perceived as more painful than paresthesia pain that feels like pins and needles or tingling [12]. Paroxysmal NP is common in patients with post-surgical pain or CRPS, therefore a modest improvement in pain is perceived as an impactful change by patients. In addition, conditions that lead to central neuropathic pain or central sensitization of pain, such as spinal cord injury, brain injury, and CRPS, may be felt more intensely than a small fiber or primarily peripheral neuropathy from diabetes mellitus or sarcoidosis. We observed that the Central Nervous System, Surgical, and CRPS etiologies had the smallest average MCIDs, which may reflect the possibility that they have an attenuated score range. Conversely, patients with DPN are likely to experience paresthesia’s, meaning a more pronounced improvement in pain may be required before it is recognized as meaningful. This may generate greater variability and an MCID that may be larger than other etiologies. The NPSI contains 5 subscales measuring specific symptom categories in NP including burning pain, pressing pain, paroxysmal pain, evoked pain, and paresthesia. Evaluating these NPSI subscales to characterize pain for each NP etiology may confirm these explanations. For this meta-analysis, NPSI sub-scales were too infrequently reported to calculate individual MCIDs. Other factors may also contribute to variation in MCID. Patients with chronic conditions are likely to experience fluctuations in daily pain levels, which introduces a higher level of variance in the sample, whereas post-surgical pain conditions are likely to improve more gradually over time, however the timeframes of recovery vary. Etiology specific MCID values would increase the precision of MCID estimates in the context of specific sub-populations of NP. However, the etiology specific results presented here must be interpreted with caution. All of the etiology estimates had a very low within group number of treatments arm. This is reflected in the large prediction intervals for etiology categories observed in both of the meta-regression analyses. These factors create uncertainty that etiology specific MCID estimates vary due to characteristics of the NP subtype or due to random sampling error. Before reliable etiology specific MCID values for the NPSI can be understood, more studies will need to be completed to add to the evidence base.

The findings of this meta-analysis are pertinent for implementation of the NPSI in clinical care and to facilitate the interpretation of clinical outcomes. Although the NPSI is a validated assessment tool [11], a substantial number of screened studies used the NPSI to characterize pain. While this suggests additional utility of the NPSI as a screening tool for classification, the NPSI is underutilized as a primary outcome to quantify changes in neuropathic pain. The value of the NPSI to discriminate distinct types of neuropathic pain for clinical evaluation is accepted, however the NPSI is also advantageous for monitoring neuropathic pain as it is validated and responsive to changes in each pain domain [40].

Future research can improve the understanding of the MCID of the NPSI by publishing anchor-based estimates based on their trial data [20]. In addition, as more trials are published, distributionally and anchor-based estimates of MCID can be calculated that are specific to interventions and etiologies.

As the NPSI continues to be used as a metric of improvement in intervention trials for NP, understanding the MCID of this scale will become increasingly important. This meta-analysis produced distribution-based MCID estimates from currently available published intervention trials. A limitation of this work is that since there are so few published trials using the NPSI, each etiology only had a few trial arms to base a MCID upon. In order to estimate a general MCID for the NPSI, it was necessary to aggregate across etiologies. However, treatment effects can be modulated within an etiology and thus future targeted MCID estimates based on large samples of data would allow for greater precision in understanding the magnitude of clinical improvement within NP subtypes. While this analysis included a highly heterogenous set of interventions, this is not a limitation, as MCID values are not specific to an intervention, but rather to the scale itself. This analysis succeeds at producing an initial broad estimate of the MCID for the NPSI within NP. The results of this analysis are more relevant to the properties of the NPSI that indicate the clinically important magnitude of improvement on this PRO for NP in general, and are less relevant for understanding the MCID of this scale for a specific NP etiology.

In conclusion, this meta-analysis has used distributional methods to produce estimates of the MCID for the NPSI total that are general across the examined etiologies and interventions. These estimates can be used to guide clinical trial design or to better understand the clinical importance of published NPSI total results.

## Data Availability

All relevant data are within the manuscript and/or its Supporting Information files.

## Acknowledgements

Dr. Jeffrey Bower is a paid employee of Sana Health, Inc. There are no conflicts of interest with any of the authors of this study. No sources of financial or other support to declare.

Additional material including analytic code, data extracted from individual studies, data for excluded studies, and quality analysis are available from the authors upon reasonable request. This protocol was developed in accordance with PRISMA guidelines and registered on PROSEPRO (CRD42025649343)

## References

1. Baron, R., Binder, A., & Wasner, G. (2010). Neuropathic pain: diagnosis, pathophysiological mechanisms, and treatment. The Lancet Neurology, 9(8), 807–819

2. GBD 2016 Disease and Injury Incidence and Prevalence Collaborators. (2017). Global, regional, and national incidence, prevalence, and years lived with disability for 328 diseases and injuries for 195 countries, 1990–2016: a systematic analysis for the Global Burden of Disease Study 2016. The Lancet, 390(10100), 1211–1259

3. Sadosky, A., McDermott, A. M., Brandenburg, N. A., & Strauss, M. (2013). A review of the epidemiology of painful diabetic peripheral neuropathy, postherpetic neuralgia, and less commonly studied neuropathic pain conditions. Pain Practice, 14(1), S56–S67. 10.1111/papr.12050

4. van Hecke, O., Austin, S. K., Khan, R. A., Smith, B. H., & Torrance, N. (2014). Neuropathic pain in the general population: a systematic review of epidemiological studies. Pain, 155(4), 654–662

5. Treede, R.D., Rief, W., Barke, A., Aziz, Q., Bennett, M.I., Benoliel, R., Cohen, M., Evers, S., Finnerup, N.B., First, M.B. & Giamberardino, M.A., (2019). Chronic pain as a symptom or a disease: the IASP Classification of Chronic Pain for the International Classification of Diseases (ICD-11). pain, 160(1), 19–27

6. Scholz, J., Finnerup, N.B., Attal, N., Aziz, Q., Baron, R., Bennett, M.I., Benoliel, R., Cohen, M., Cruccu, G., Davis, K.D. & Evers, S., (2019). The IASP classification of chronic pain for ICD-11: chronic neuropathic pain. PAIN, 160, 53–59.

7. Todorovic, M.S., Frey, K., Swarm, R.A., Bottros, M., Rao, L., Tallchief, D., Kraus, K., Meacham, K., Bakos, K., Zang, X. & Lee, J.B., (2022). Prediction of individual analgesic response to intravenous lidocaine in painful diabetic peripheral neuropathy: a randomized, placebo-controlled, crossover trial. The Clinical journal of pain, 38(2), 65–76.

8. Ushida, T., Katayama, Y., Hiasa, Y., Nishihara, M., Tajima, F., Katoh, S., Tanaka, H., Maeda, T., Furusawa, K., Richardson, M. & Kakehi, Y., (2023). Mirogabalin for central neuropathic pain after spinal cord injury: a randomized, double-blind, placebo-controlled, phase 3 study in Asia. Neurology, 100(11), e1193–e1206.

9. Wong, M. L., Widerstrom-Noga, E., & Field-Fote, E. C. (2022). Effects of whole-body vibration on neuropathic pain and the relationship between pain and spasticity in persons with spinal cord injury. Spinal cord, 60(11), 963–970.

10. Zhang, Y., Xu, L., Huang, Y. (2024). Update in the Treatment of Neuropathic Pain. In: Ma, C., Huang, Y. (eds) Translational Research in Pain and Itch. Springer, Singapore. 10.1007/978-981-99-8921-8_12

11. Attal, N., Bouhassira, D., & Baron, R. (2018). Diagnosis and assessment of neuropathic pain through questionnaires. The Lancet Neurology, 17(5), 456–466.

12. Backonja, M.M. & Stacey B. (2004). Neuropathic pain symptoms relative to overall pain rating. J Pain, 5(9), 491–497.

13. Haanpää, M. L., & Treede, R. D. (2010). Diagnosis and classification of neuropathic pain. Pain, 150(1), 14–27

14. Bouhassira, D., Attal, N., Alchaar, H., Boureau, F., Brochet, B., Bruxelle, J., Cunin, G., Fermanian, J., Ginies, P., Grun-Overdyking, A., Jafari-Schluep, H., Lantéri-Minet, M., Laurent, B., Mick, G., Serrie, A., Valade, D., & Vicaut, E. (2005). Comparison of pain syndromes associated with nervous or somatic lesions and development of a new neuropathic pain diagnostic questionnaire (DN4). Pain, 114(1-2), 29–36

15. Haanpää, M. L., Attal, N., Backonja, M., Baron, R., Bennett, M., Bouhassira, D., … & Treede, R. D. (2011). NeuPSIG guidelines on neuropathic pain assessment. PAIN®, 152(1), 14–27.

16. U.S. Food and Drug Administration. (2022, January 26). Principles for selecting, developing, modifying, and adapting patient-reported outcome instruments for use in medical device evaluation: Guidance for industry and Food and Drug Administration staff, and other stakeholders. https://www.fda.gov/media/141565/download

17. Jaeschke, R., Singer, J., & Guyatt, G. H. (1989). Measurement of health status: ascertaining the minimal clinically important difference. Controlled clinical trials, 10(4), 407–415.

18. Fritz, C. O., Morris, P. E., & Richler, J. J. (2012). Effect size estimates: Current use, calculations, and interpretation. Journal of Experimental Psychology: General, 141(1), 2–18. 10.1037/a0024338

19. Howard, R., Phillips, P., Johnson, T., O’Brien, J., Sheehan, B., Lindesay, J., Bentham, P., Burns, A., Ballard, C., Holmes, C. & McKeith, I., (2011). Determining the minimum clinically important differences for outcomes in the DOMINO trial. International journal of geriatric psychiatry, 26(8), 812–817.

20. Copay, A. G., Subach, B. R., Glassman, S. D., Polly Jr, D. W., & Schuler, T. C. (2007). Understanding the minimum clinically important difference: a review of concepts and methods. The Spine Journal, 7(5), 541–546.

21. Furukawa, T. A., Barbui, C., Cipriani, A., Brambilla, P., & Watanabe, N. (2006). Imputing missing standard deviations in meta-analyses can provide accurate results. Journal of clinical epidemiology, 59(1), 7–10.

22. McLeod, L. D., Coon, C. D., Martin, S. A., Fehnel, S. E., & Hays, R. D. (2011). Interpreting patient-reported outcome results: US FDA guidance and emerging methods. Expert review of pharmacoeconomics & outcomes research, 11(2), 163–169.

23. U.S. Food and Drug Administration. (2009). Patient-reported outcome measures: Use in medical product development to support labeling claims. U.S. Department of Health and Human Services. Retrieved from https://www.fda.gov/regulatory-information/search-fda-guidance-documents/patient-reported-outcome-measures-use-medical-product-development-support-labeling-claims

24. Norman GR, Sloan JA, Wyrwich KW. Interpretation of changes in health-related quality of life. The remarkable universality of half a standard deviation. Med Care 2003;41:582–92

25. Higgins, J. P. T., Altman, D. G., & Sterne, J. A. C. (Eds.). (2011). Chapter 8: Assessing risk of bias in included studies. In J. P. T. Higgins & S. Green (Eds.), Cochrane handbook for systematic reviews of interventions (Version 5.1.0). The Cochrane Collaboration. Available from https://handbook-5-1.cochrane.org

26. de Oliveira Rocha, R., Teixeira, M.J., Yeng, L.T., Cantara, M.G., Faria, V.G., Liggieri, V., Loduca, A., Müller, B.M., Souza, A.C. & de Andrade, D.C., (2014). Thoracic sympathetic block for the treatment of complex regional pain syndrome type I: a double-blind randomized controlled study. PAIN®, 155(11), 2274–2281.

27. Hoerder, S., Habermann, I.V., Hahn, K., Meyer-Hamme, G., Ortiz, M., Grabowska, W., Roll, S., Willich, S.N., Schroeder, S., Brinkhaus, B. & Dietzel, J. (2023). Acupuncture in diabetic peripheral neuropathy-neurological outcomes of the randomized acupuncture in diabetic peripheral neuropathy trial. World journal of diabetes, 14(12), 1813.

28. Schlaeger, J. M., Molokie, R. E., Yao, Y., Suarez, M. L., Golembiewski, J., Wilkie, D. J., & Votta-Velis, G. (2017). Management of sickle cell pain using pregabalin: a pilot study. Pain Management Nursing, 18(6), 391–400.

29. Culver, D.A., Dahan, A., Bajorunas, D., Jeziorska, M., van Velzen, M., Aarts, L.P., Tavee, J., Tannemaat, M.R., Dunne, A.N., Kirk, R.I. & Petropoulos, I.N., (2017). Cibinetide improves corneal nerve fiber abundance in patients with sarcoidosis-associated small nerve fiber loss and neuropathic pain. Investigative ophthalmology & visual science, 58(6), BIO52–BIO60.

30. Rehm, S., Binder, A., & Baron, R. (2010). Post-herpetic neuralgia: 5% lidocaine medicated plaster, pregabalin, or a combination of both? A randomized, open, clinical effectiveness study. Current medical research and opinion, 26(7), 1607–1619.

31. Raksakietisak, M., Rushatamukayanunt, P., Wilaiwan, K., Homprasert, C., Nitising, A., Sawasdiwipachai, P., & Pantubtim, C. (2022). Postoperative analgesia of intraoperative nefopam in patients undergoing anterior cervical spine surgery: A prospective randomized controlled trial. Medicine, 101(43), e31296.

32. Wan, X., Wang, W., Liu, J., & Tong, T. (2014). Estimating the sample mean and standard deviation from the sample size, median, range and/or interquartile range. BMC Medical Research Methodology, 14(1), 135. 10.1186/1471-2288-14-135

33. Juhl, A. A., Karlsson, P., & Damsgaard, T. E. (2016). Fat grafting for alleviating persistent pain after breast cancer treatment: a randomized controlled trial. Journal of Plastic, Reconstructive & Aesthetic Surgery, 69(9), 1192–1202.

34. Martinez, V., Szekely, B., Lemarié, J., Martin, F., Gentili, M., Ammar, S.B., Lepeintre, J.F., de Loubresse, C.G., Chauvin, M., Bouhassira, D. & Fletcher, D., (2013). The efficacy of a glial inhibitor, minocycline, for preventing persistent pain after lumbar discectomy: a randomized, double-blind, controlled study. PAIN®, 154(8), 1197–1203.

35. Kim, J. K., Park, H. S., Bae, J. S., Jeong, Y. S., Jung, K. J., & Lim, J. Y. (2020). Effects of multi-session intermittent theta burst stimulation on central neuropathic pain: a randomized controlled trial. NeuroRehabilitation, 46(1), 127–134.

36. Redelmeier, D. A., & Kahneman, D. (1996). Patients’ memories of painful medical treatments: Real-time and retrospective evaluations of two minimally invasive procedures. pain, 66(1), 3–8.

37. R Core Team (2023). _R: A Language and Environment for Statistical Computing_. R Foundation for Statistical Computing, Vienna, Austria. <https://www.R-project.org/>.

38. Viechtbauer, W. (2010). Conducting meta-analyses in R with the metafor package. Journal of Statistical Software, 36(3), 1–48. 10.18637/jss.v036.i03

39. Watt, J. A., Veroniki, A. A., Tricco, A. C., & Straus, S. E. (2021). Using a distribution-based approach and systematic review methods to derive minimum clinically important differences. BMC Medical Research Methodology, 21, 1–7.

40. Lucchetta, M., Pazzaglia, C., Padua, L., & Briani, C. (2011). Exploring neuropathic symptoms in a large cohort of Italian patients with different peripheral nervous system diseases. Neurological Sciences, 32(3), 423–426.

